# Epidemiological and genomic investigation of chikungunya virus in Rio de Janeiro state, Brazil, between 2015 and 2018

**DOI:** 10.1101/2023.04.12.23288482

**Authors:** Filipe Romero Rebello Moreira, Mariane Talon de Menezes, Clarisse Salgado-Benvindo, Charles Whittaker, Victoria Cox, Nilani Chandradeva, Hury Hellen Souza de Paula, André Frederico Martins, Raphael Rangel das Chagas, Rodrigo Decembrino Vargas Brasil, Darlan da Silva Cândido, Alice Laschuk Herlinger, Marisa de Oliveira Ribeiro, Monica Barcellos Arruda, Patricia Alvarez, Marcelo Calado de Paula Tôrres, Ilaria Dorigatti, Oliver Brady, Carolina Moreira Voloch, Amilcar Tanuri, Felipe Iani, William Marciel de Souza, Sergian Vianna Cardozo, Nuno Rodrigues Faria, Renato Santana de Aguiar

## Abstract

Since 2014, Brazil has experienced an unprecedented epidemic caused by chikungunya virus (CHIKV), with several waves of East-Central-South-African (ECSA) lineage transmission reported across the country. In 2018, Rio de Janeiro state, the third most populous state in Brazil, reported 41% of all chikungunya cases in the country. Here we use evolutionary and epidemiological analysis to estimate the timescale of CHIKV-ECSA-American lineage and its epidemiological patterns in Rio de Janeiro. We show that the CHIKV-ECSA outbreak in Rio de Janeiro derived from two distinct clades introduced from the Northeast region in mid-2015 (clade RJ1, *n* = 63/67 genomes from Rio de Janeiro) and mid-2017 (clade RJ2, *n* = 4/67). We detected evidence for positive selection in non-structural proteins linked with viral replication in the RJ1 clade (clade-defining: nsP4-A481D) and the RJ2 clade (nsP1-D351G). Finally, we estimate the CHIKV-ECSA’s basic reproduction number (*R_0_*) to be between 1.2 to 1.6 and show that its instantaneous reproduction number (*R_t_*) displays a strong seasonal pattern with peaks in transmission coinciding with periods of high *Aedes aegypti* transmission potential. Our results highlight the need for continued genomic and epidemiological surveillance of CHIKV in Brazil, particularly during periods of high ecological suitability, and show that selective pressures underline the emergence and evolution of the large urban CHIKV-ECSA outbreak in Rio de Janeiro.

## 1. Introduction

Chikungunya virus (CHIKV) is an enveloped virus with a single-stranded positive-sense RNA 11.8 kb genome that belongs to the *Alphavirus* genus in the *Togaviridae* family [1]. The CHIKV genome contains two open reading frames encoding four non-structural proteins (nsP1–nsP4), important for viral replication, and four structural proteins (capsid, E1–E3), necessary for virus assembly [2]. CHIKV is transmitted by the bites of infected *Aedes* spp. mosquitoes, mainly from the *Ae*. *aegypti* species and, to a lesser extent, the *Ae*. *albopictus* species. In humans, clinical manifestations of chikungunya include arthralgia, high fever, myalgia, headache and often exanthema. However, chikungunya may also cause long-lasting debilitating polyarthralgia [1], and can also lead to neurological complications and fatal outcomes [3]. Despite being an important public health threat with >1 billion people at risk for transmission [4], reporting of chikungunya infections often relies on syndromic surveillance, which is challenged by the co-circulation of mosquito-borne viruses that cause similar clinical symptoms, including dengue (DENV) and Zika (ZIKV) [5].

Since its first description in Tanzania in 1952, chikungunya has caused over 70 outbreaks in Africa, Asia, Americas, Europe and in Ocean Pacific Islands [4]. CHIKV can be classified into four distinct lineages: the West-African lineage, the Asian lineage, the East-Central-South-African (ECSA) genotype [6], and the Indian Ocean lineage (IOL). Chikungunya’s geographic distribution – particularly of Asian, IOL and ECSA lineages – has been expanding rapidly over the last 20 years probably due rapid urbanization, globalization of trade, and virus evolution and adaptation to local variation in the distribution of vector species [7]. For example, a single amino acid change in the E1 protein (*i.e.*, E1-A226V) of the CHIKV IOL lineage has been linked to increase transmissibility and infectivity in *Ae*. *albopictus* and was linked with a series of severe outbreaks in Indo-Pacific Asia [8–10].

In 2013, the CHIKV Asian lineage was first detected in St Martin islands in the Caribbean [11]. Since then, this lineage has spread to >50 countries and territories in the Americas [11, 12]. In 2014, co-circulation of the Asian and ECSA lineages was detected in Brazil [13]. Local transmission of the Asian genotype was first detected in Amapá state, North Brazil, on 31 July 2014 [13]. On 25 August 2014, local transmission of the CHIKV ECSA lineage was identified in Bahia state, Northeast Brazil [13, 14]. Retrospective outbreak investigations revealed that the ECSA index case in Brazil arrived in Bahia in late May 2014 from Angola [14]. This new ECSA-American lineage was then linked to large outbreaks in the Amazonas and Roraima states in North Brazil, despite earlier circulation of the Asian lineage in that region [15]. Currently, circulation of ECSA-American lineage has been confirmed in all five geographic regions of Brazil [15–20], but also in Paraguay [21] and Haiti [22].

Rio de Janeiro (RJ) is the third most populous Brazilian state and a key business and touristic hub in the Americas. Due to air and fluvial connectivity and high mosquito climatic suitability [23], the state has historically suffered large mosquito-borne viral outbreaks [24], including dengue, Zika, yellow fever and, more recently, chikungunya. Between 2013 and 2018, CHIKV has caused 403,828 confirmed infections in Brazil [25]. In 2018, RJ became the epidemic center of CHIKV in Brazil, accounting for 41% of the cases reported in the country [25]. Until 2017, only 20,251 cases had been confirmed in Rio de Janeiro, corresponding to 6% of the total case numbers reported in Brazil. The earliest autochthonous chikungunya cases were reported in November 2015, and since then two ECSA sub-lineages have been detected in Rio de Janeiro [17, 18, 26]. Despite the burden imposed in Rio de Janeiro, chikungunya’s evolution and transmission patterns remain poorly understood. The reliance of chikungunya diagnosis on clinical-epidemiological criteria and the scarcity of chikungunya sequence data from Rio de Janeiro, hamper our understanding of chikungunya transmission and evolution. Moreover, relevant epidemiological parameters, such as basic (R_0_) and time varying (R_t_) reproduction numbers, which measure virus transmissibility, have not been yet previously estimated for CHIKV in Rio de Janeiro, hindering comparisons with the virus’ epidemiological dynamics in other settings.

Using genetic analysis of CHIKV newly generated and publicly available genome sequences and CHIKV traditional surveillance data, we investigated its evolution and transmission dynamics in Rio de Janeiro. We show that the large 2018 CHIKV outbreak in Rio de Janeiro was mainly caused by one dominant ECSA-American viral clade that likely persisted locally since mid-2015. In addition, we provide evidence of adaptive molecular evolution on non-structural amino acid positions, including in a lineage-defining mutation related with the dominant ECSA viral clade associated with the 2018 outbreak in Rio de Janeiro. Finally, using epidemiological modeling, we estimate basic and instantaneous reproduction numbers for CHIKV ECSA-American in Brazil, further expanding our understanding of the epidemiology of this rapidly expanding lineage.

## 2. Materials and Methods

### 2.1. Quantifying CHIKV transmissibility in Rio de Janeiro

The weekly aggregated epidemiological data of chikungunya laboratory-confirmed cases in RJ was obtained from the Brazilian Ministry of Health from 2014 to 2018, and then the results were presented in time series. Data was used to estimate weekly CHIKV incidence and transmissibility (reproduction number) for Rio de Janeiro between 2016 and 2018. We estimated the basic reproductive number (*R_0_*), defined as the number of secondary infections caused by an infected individual in a completely susceptible population, using data from 3 January to 13 March of 2016. We employed the exponential growth method to estimate *R_0_* [27], which derives an estimate of *R_0_* from the generation time distribution along with an estimate of the exponential growth rate, calculated during the earliest weeks of the outbreak where epidemic growth was approximately exponential and factors such as susceptible depletion, behavioral change or vector control are negligible. To assess changes in transmission over time in subsequent epidemics when immunity due to prior natural infection is present, we also calculated the instantaneous reproduction number (*R_t_*), defined as the average number of secondary cases caused by an infected individual during the epidemic, provided the conditions remained as they were at time *t* [28]. We used an 8-week sliding-window to estimate R_t_, and conducted several sensitivity analyses considering different sliding-window intervals (3 to 7 weeks). We assumed a parametric gamma distribution for the generation time (GT) with a mean of 14 days and a standard deviation of 6.4 days, based on an earlier estimate obtained during an outbreak caused by the Indian Ocean lineage [29]. We also performed sensitivity analyses with different mean GT values (10 and 20 days), keeping the standard deviation constant. All analyses were performed in R software v4.2.2 [30] with the *R_0_* [31] and EpiEstim [28] packages.

### 2.2. *Aedes aegypti* transmission potential in Rio de Janeiro

To estimate *Aedes aegypti* transmission potential for RJ between 2014 and 2018, we first extracted daily values (midday) of 2m air temperature, 2m dew point temperature and total precipitation for all locations across RJ from the ERA-5 ECMWF reanalysis product [32]. We calculated relative humidity based on daily temperature and dew point estimates using the August-Roche-Magnus approximation [33–35]. Each variable was then averaged over RJ and combined to calculate index P using the MVSE R package [36]. We used priors related to *Ae. aegypti* ecology and entomology, as described elsewhere [36]. We then aggregated resulting daily estimates to generate mean weekly estimates for index P between 2014 and 2018. Code and data used in these analyses are available in **Supplementary File S1**.

### 2.3. Sample collection, RNA extraction, and CHIKV molecular diagnosis

To assess the genetic diversity of CHIKV in Rio de Janeiro state, we generated new CHIKV genomes obtained from samples collected in the Duque de Caxias municipality, the third most populous city in Rio de Janeiro state (**Figures 1A, 1B**; https://cidades.ibge.gov.br/brasil/rj/duque-de-caxias/, last accessed 21 June, 2022). Samples were collected following approval of an ethical review board (CAAE 54544316.3.0000.5283). A total of 179 samples, blood or urine, from arboviral suspected cases in public health care units between July 2017 and June 2018. Viral nucleic acid extractions were performed with the QIAmp viral RNA mini kit (Qiagen, Germany), following manufacturer’s instructions. We then tested extracted RNAs by RT-qPCR for detection of ZIKV, DENV, and CHIKV using the ZDC molecular kit (Bio-Manguinhos FIOCRUZ, Brazil). Samples presenting a cycle threshold (Ct) < 38 for a given target were considered positive for CHIKV.

**Figure 1:**
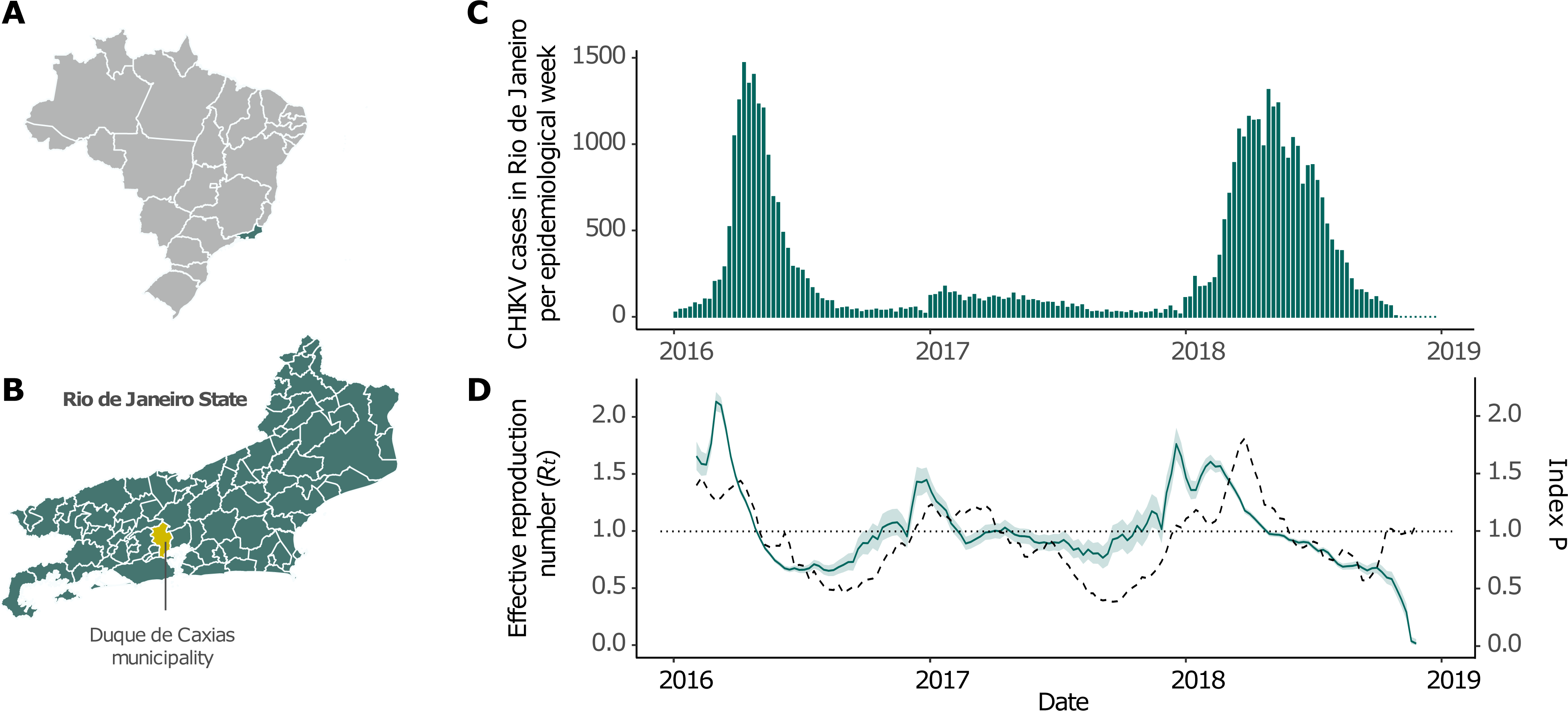
Map of Brazil and Rio de Janeiro state, state-level weekly CHIKV incidence in the 2016-2018 period and *R_t_* estimate. **(A)** Map of Brazil with all states colored in light gray, except for Rio de Janeiro, in green. **(B)** Rio de Janeiro map with municipality borders delimitation. The municipality of Duque de Caxias, where samples for genome sequencing were collected, is colored yellow. **(C)** State level weekly incidence data. Bars correspond to the number of CHIKV cases. **(D)** Estimate for *Rt* between 2016 and 2018 (left y-axis). The solid line indicates mean values, while the ribbon indicates the 95% confidence interval. The dashed line represents index P values, a measure of transmissibility potential for the vector *Aedes aegypti* (right y-axis). The dotted line marks the critical epidemic threshold (*R_t_* = 1).

### 2.4. Chikungunya virus whole-genome sequencing

We selected 34 CHIKV RNA-positive samples with Ct < 30 for viral whole-genome sequencing using the Illumina platform. Samples were distributed across the most sampled months of 2018 (March, April, and May), and one sample from 2017 was also sequenced. Amplicon target sequencing was conducted employing an amplicon-based sequencing protocol [37]. Briefly, we converted viral RNA to cDNA with SuperScript IV (Thermo Fisher Scientific), which was amplified with the Q5 DNA polymerase (New England Biolabs, UK) in two PCR reactions. Each PCR reaction contained 22 pairs of primers and generated overlapping amplicons of approximately 400 bp [37]. The amplicons covered the entire CHIKV genome and were used as input to the QIAseq 1-Step Library prep kit (Qiagen, Germany), following the manufacturer’s protocol. Libraries were normalized and equimolarly pooled at 14 pMol with 10% phiX control. Sequencing was performed on an Illumina MiSeq instrument with a V3 (600 cycles) cartridge.

### 2.5. Virus genome assembly

Strict quality control of raw sequencing reads was performed with Trimmomatic v.0.39 [38], which removed low-quality bases (Phred scores < 30), sequencing adapters, and short reads (<50 nucleotides). This software was also used to conservatively trim the 30 initial nucleotides from all reads, eliminating primer-related sequences. Filtered reads from each sample were then mapped against a reference CHIKV genome (NCBI accession number: KP164568.1) with Bowtie2 v2.4.2 [39]. Mapping files were sorted and indexed with samtools v1.11 [40]. Bcftools v1.11 [41] was used to call variants with a high-quality threshold (QUAL>200) and to estimate consensus genome sequences. Finally, bedtools v2.30.0 [42] was used to mask low-coverage sites (<10x). This assembly pipeline is publicly available on GitHub (https://github.com/filiperomero2/ViralUnity, last accessed 20 June 2022). The novel consensus genome sequences belonged to the ECSA lineage and were deposited to NCBI GenBank under Accession Numbers OP312936 to OP312969 (**Table 1**).

**Table 1.** Sample metadata and sequencing statistics for Duque de Caxias samples.

| **Sample ID** | **NCBI accession** | **Collection date** | **RT-qPCR Ct** | **Number of reads** | **Average depth** | **Genome coverage (%)** |
| --- | --- | --- | --- | --- | --- | --- |
| 10 | OP312936 | 2017-07-26 | 23.55 | 634,418 | 2582.12 | 85.84 |
| 53 | OP312937 | 2018-03-01 | 19.45 | 738,782 | 4041.52 | 97.24 |
| 57 | OP312938 | 2018-03-08 | 18.97 | 1,311,212 | 10039 | 98.00 |
| 66 | OP312939 | 2018-03-15 | 14.73 | 624,422 | 3281.5 | 94.34 |
| 71 | OP312940 | 2018-03-19 | 18.78 | 900,426 | 3051.64 | 70.50 |
| 79 | OP312941 | 2018-03-22 | 17.08 | 924,090 | 4257.63 | 95.15 |
| 83 | OP312942 | 2018-03-27 | 19.84 | 565,152 | 4016.5 | 96.19 |
| 89 | OP312943 | 2018-03-29 | 20.08 | 1,520,304 | 7710.58 | 94.39 |
| 92 | OP312944 | 2018-03-29 | 19.19 | 1,009,462 | 5340.61 | 97.68 |
| 97 | OP312945 | 2018-04-02 | 19.35 | 1,159,296 | 5811.29 | 96.74 |
| 99 | OP312946 | 2018-04-02 | 19.91 | 2,310,894 | 5509.86 | 84.85 |
| 101 | OP312947 | 2018-04-02 | 24.94 | 6,875,476 | 45036.2 | 95.21 |
| 107 | OP312948 | 2018-04-04 | 19.57 | 841,146 | 2703.55 | 84.18 |
| 110 | OP312949 | 2018-04-05 | 20.46 | 1,365,558 | 3543.14 | 78.29 |
| 114 | OP312950 | 2018-04-05 | 20.04 | 1,557,664 | 6901.02 | 93.02 |
| 116 | OP312951 | 2018-04-05 | 19.62 | 663,888 | 3471.67 | 93.19 |
| 117 | OP312952 | 2018-04-05 | 23.45 | 876,300 | 5417.87 | 90.16 |
| 120 | OP312953 | 2018-04-16 | 18.07 | 765,232 | 2285.81 | 94.73 |
| 126 | OP312954 | 2018-04-17 | 23.02 | 1,405,840 | 6905.76 | 87.42 |
| 128 | OP312955 | 2018-04-19 | 21.72 | 1,365,992 | 4532.91 | 84.24 |
| 130 | OP312956 | 2018-04-19 | 18.47 | 1,038,668 | 4913.46 | 94.99 |
| 134 | OP312957 | 2018-04-19 | 22.77 | 2,133,788 | 10443.2 | 94.98 |
| 143 | OP312958 | 2018-04-30 | 23.22 | 1,518,024 | 7322.5 | 91.43 |
| 149 | OP312959 | 2018-04-30 | 23.77 | 1,034,102 | 1806.96 | 85.10 |
| 152 | OP312960 | 2018-05-03 | 24.85 | 2,261,360 | 9137.28 | 87.06 |
| 155 | OP312961 | 2018-05-03 | 24.84 | 1,339,770 | 6698.01 | 85.11 |
| 157 | OP312962 | 2018-05-07 | 21.03 | 1,778,388 | 8387.42 | 89.23 |
| 159 | OP312963 | 2018-05-07 | 24.37 | 628,560 | 2628.08 | 90.56 |
| 161 | OP312964 | 2018-05-07 | 24.73 | 1,182,780 | 3850.23 | 89.45 |
| 165 | OP312965 | 2018-05-09 | 18.83 | 1,150,356 | 6482.64 | 96.64 |
| 166 | OP312966 | 2018-05-09 | 22.01 | 1,467,544 | 7224.4 | 86.22 |
| 170 | OP312967 | 2018-05-14 | 24.08 | 1,048,464 | 4201.88 | 91.86 |
| 174 | OP312968 | 2018-05-17 | 14.76 | 1,092,190 | 4108.37 | 95.44 |
| 181 | OP312969 | 2018-05-23 | 18.75 | 816,186 | 4507.98 | 97.82 |

### 2.6. Maximum likelihood and Bayesian phylogenetic analyses

To contextualize the novel genome sequences within the global CHIKV diversity, we downloaded all CHIKV sequences with more than 8,000 nucleotides available on NCBI GenBank [43], as of 31 May 2022. Sequences without information on the country of sampling and date of collection were removed from subsequent analyses. The remaining 1,262 sequences were combined with the 34 new sequences and aligned to the CHIKV RefSeq genome (NCBI accession: NC_004162.2) with MAFFT v7.480 [44]. After manually trimming gap-rich regions, we estimated maximum-likelihood phylogenetic trees using IQ-Tree v2.1.2 [45]. We used Model-Finder [46] to determine the best-fit substitution model and the Shimoidara-Hasegawa-like approximate likelihood ratio test (SH-aLRT) [47] to assess phylogenetic node support.

ECSA-American sub-lineage sequences from countries outside of Brazil were excluded from subsequent analyses, and maximum likelihood phylogenetic analyses were performed separately with ECSA sequences from Brazil collected up to the end of 2018 using the methods described above. Sequences were aligned against the oldest available ECSA strain from Brazil (Accession number: KP164568, from Feira de Santana, Bahia state), and non-coding regions were trimmed out of the alignment. Screening of recombination signals was performed using RDP4 [48].

### 2.7. Temporal structure of CHIKV ECSA-American nucleotide alignment

Root-to-tip regressions were used to evaluate the temporal signal available on genomes from the ECSA-American sub-lineage dataset, including only Brazilian strains. The estimated maximum likelihood phylogenetic tree was manually rooted in the oldest available ECSA strain from Brazil and regression between divergence and sampling dates was analyzed in TempEst v1.5.3 [49]. To optimize the temporal signal, outliers were conservatively defined as sequences whose regression residuals exceeded more or less than two times the interquartile range of the residual distribution, and 51 sequences were removed. A maximum likelihood tree inference and root-to-tip regression were performed on the Brazilian ECSA-American sub-lineage (*n* = 148).

### 2.8. Bayesian phylogenetic analyses

To investigate the evolutionary origins of CHIKV in Rio de Janeiro, we used Bayesian coalescent and phylogeographic models available in BEAST v1.10.4 [50, 51]. Analyses were run using a HKY+G4 nucleotide substitution model [52, 53], a strict molecular clock model, and a flexible skygrid tree prior [54]. The cut-off for the skygrid model was set at 4.22 years, following a preliminary estimate of the time of the most recent common ancestor (tMRCA) obtained from TempEst (x-axis intercept in the regression). The number of grid points was set to match the number of months within the tree temporal span between the tMRCA and the earliest tip (*n* = 51). We used default priors and operators, and ran two independent Markov Chain Monte Carlo (MCMC) chains with 100 million steps, sampling every 10,000th step, using a maximum-likelihood phylogeny as a starting tree. The BEAGLE library was used for accelerated computations [55]. We assessed mixing of MCMC chains and convergence of all parameters using Tracer v1.7 [56]. We used Logcombiner [50] to sample 1,000 trees from the combined posterior trees distribution, after removal of 10% burn-in. A discrete phylogeographic analysis was then performed with a set of 1,000 empirical dated trees obtained from the posterior distribution, as previously described [57]. We used an asymmetric substitution model [51] and considered four geographic locations/regions: Rio de Janeiro state (*n* = 67), North region (*n* = 22), Northeast region (*n* = 52), and Central-West region (*n* = 7).

Finally, we also investigated changes in CHIKV genetic diversity over time in RJ. We assembled a dataset comprehending all Rio de Janeiro sequences belonging to the dominant clade (*n* = 63) and used a skygrid prior, as described above. In this case, we used an informative prior on the root age based on the estimate obtained from the analysis of the full dataset. BEAST analyses were run in duplicate for 20 million generations, sampling every 2,000^th^ state. BEAST XML files and outputs are available in **Supplementary File S2**.

### 2.9. Selection analyses and ancestral states reconstruction

Selection analyses were conducted on the ECSA-American alignment with Brazilian sequences described above using MEME (Mixed Effects Model of Evolution) [58] and FEL (fixed effects likelihood) [59] models to detect individual genome sites subjected to episodic and pervasive positive selection on the Datamonkey web server [60, 61]. A significance threshold of *p* < 0.01 was used for analyses. To map positively selected sites along the evolutionary history of Brazilian CHIKV ECSA-American sub-lineage, we performed ancestral sequence reconstruction using TreeTime [62].

## 3. Results

### 3.1. State-level chikungunya incidence and transmissibility

The first chikungunya epidemic wave in Rio de Janeiro state occurred in 2016 (**Figure 1C**), with 16,245 laboratory-confirmed cases. In 2017, fewer cases were reported (*n* = 3,989), with a nearly constant number of infections in the first semester, decreasing by the end of the year. In early 2018, a sharp increase in chikungunya infections were observed, leading to a larger epidemic with a magnitude greater than the first (*n* = 24,990 cases). In both 2016 and 2018 waves, cases grew exponentially between January and March, reaching peaks in April. Most confirmed cases occurred in the state’s capital, Rio de Janeiro city (54%; *n* = 24,636/45,241; **Supplementary Figure S1**). This pattern was most evident in 2016 (87%; *n* = 14,204/16,245) than in 2017 (44%; *n* = 1,768/3,989) and 2018 (35%; *n* = 8,649/24,990). In 2018, while the state capital exhibited the highest incidence, a large number of infections were reported in other municipalities, such as Campos dos Goytacazes (24%; *n* = 5,968/24,990) and São Gonçalo (21%, *n* = 5,323/24,990).

To better understand the transmission dynamics of CHIKV ECSA in Rio de Janeiro state, we estimated R_0_ from notification data. First, we estimated an epidemic growth rate, *r*, of 1.032 (95% CI: 1.027 - 1.038) for 2016. Assuming a generation time (GT) of 14 days, we estimated a *R_0_* of 1.56 (95% CI = 1.46 – 1.67). Sensitivity analysis with different GT distributions generated different *R_0_* values (GT mean = 10 days, *R_0_* = 1.37, 95% CI: 1.31 - 1.44; GT mean = 20 days, *R_0_* = 1.89, 95% CI: 1.72 - 2.09) (**Supplementary Figure S2)**. Overall, these estimates are similar to those previously obtained for DENV and lower compared to *R_0_* estimates obtained for the ZIKV epidemic in Rio de Janeiro [63].

We then estimated the *Rt* to track the progress of the epidemic in Rio de Janeiro state (**Figure 1D**). Noticeably, we observed that between 2016 and mid-2018, high transmissibility periods broadly coincide with periods of relatively high *Ae. aegypti* transmission potential. This shows that during the study period, the timing of chikungunya recurrence in Rio de Janeiro was at least partly driven by climatic factors such as temperature and humidity that correlate with fluctuations in *Ae. aegypti* abundance (**Figure 1D**). As expected, *R_t_* follows a seasonal pattern, reaching the peak values in early 2016 (mean: 2.14, 95% CI: 2.06 – 2.22), 2017 (mean: 1.45, 95% CI: 1.35 – 1.56) and 2018 (mean: 1.76, 95% CI: 1.63 – 1.91). *R_t_* estimates obtained using different window lengths, and GT distributions revealed similar patterns (**Supplementary Figure S3**).

### 3.2. Molecular detection and whole-genome sequencing of CHIKV from Duque de Caxias municipality

Of the 179 clinical samples from patients presenting symptoms compatible with arbovirus infections in health-care units of the Duque de Caxias municipality, 48.6% (*n* = 87) tested positive for CHIKV RNA (mean cycle threshold, Ct, was 23.2, range: 14.7 to 33.1), 1.1% (*n* = 2) tested positive for DENV and 2.8% (*n* = 5) for ZIKV. We detected CHIKV coinfections with DENV (*n* = 1) and ZIKV (*n* = 3). For the CHIKV RNA-positive cases, clinical manifestations included fever (100 %, *n* = 87/87), arthralgia (97.7%, *n* = 85/87), headache (82.8%, *n* = 72/87), myalgia (71.3%, *n* = 62/87), arthritis (62.1%, n = 54/87), conjunctivitis (33.3%, *n* = 29/87), and exanthema (31%, *n* = 27/87) (**Figure 2A, Supplementary File S3**). Less frequently, prurience (11.5%, *n* = 10/87), dyscrasia (3.4%, *n* = 3/87) and neurological symptoms (2.3%, *n* = 2/87) were also recorded. As expected, we observed a tendency towards lower values (higher viral loads) in samples collected closer to the dates of symptom onset (**Figure 2B**). Thirty-four near complete CHIKV genome sequences were generated with an average 90.8% (standard deviation: 6.2%, range: 70.5 - 98%) genome coverage in relation to the CHIKV genome reference sequence (**Table 1**). Sequencing coverage was inversely correlated with Ct values, although no strong statistical relationship was inferred on a linear model (*p* = 0.08; **Figure 2C**).

**Figure 2:**
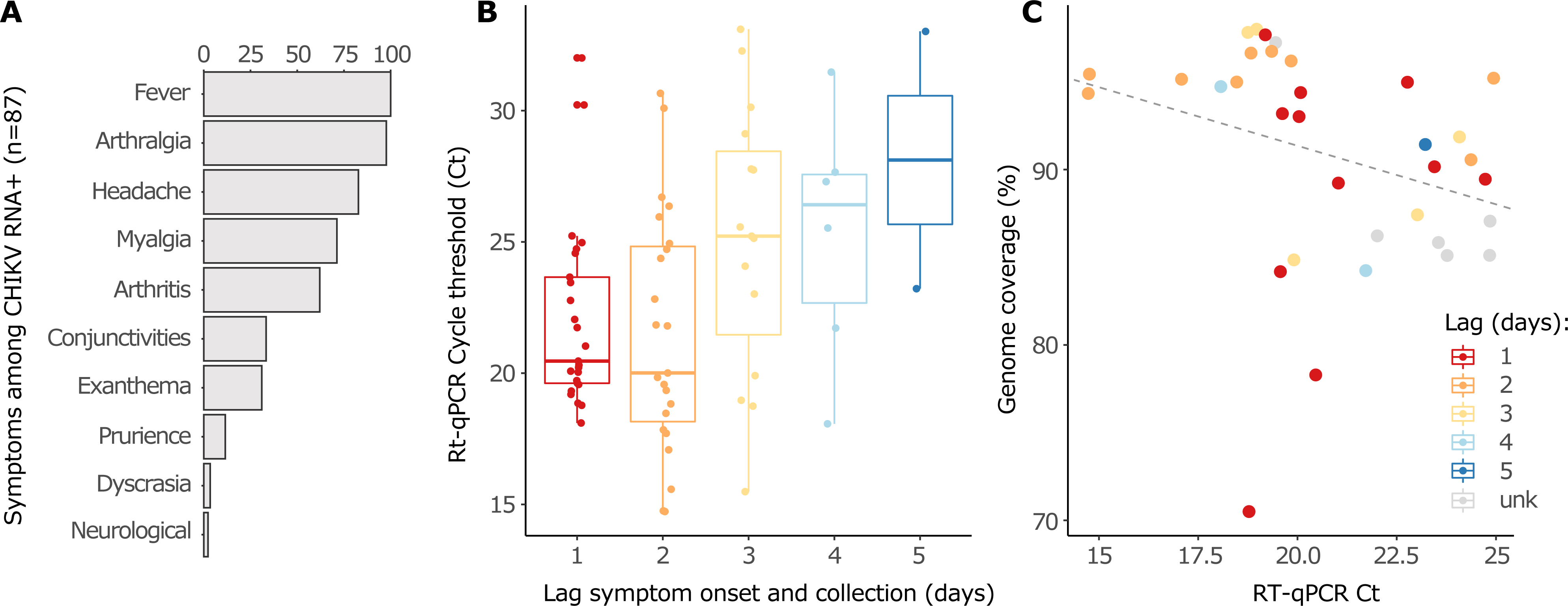
Symptoms distribution, RT-qPCR Ct values dynamics and their correlation with genome sequencing coverage. **(A)** Distribution of symptoms exhibited by all CHIKV positive patients in the Duque de Caxias cohort. **(B)** The time lag between symptoms onset and sample collection dates exhibits correlation with RT-qPCR Ct values. As infection proceeds, viral loads decrease (Cts increase) likely due to immunological response. **(C)** Negative correlation between RT-qPCR Cts and genome sequencing coverage. Sequences characterized from samples with higher viral load (lower Cts) tend to exhibit higher coverage, although no strong statistical correlation was inferred on a linear model (*p* = 0.08).

### 3.3. Phylogenetic analyses of the CHIKV ECSA-American lineage

Newly generated sequences clustered within the ECSA-American lineage with maximum statistical support (SH-aLRT = 100; **Figure 3A**). After removal of outliers, we found a strong correlation between sampling dates and genetic divergence (**Figures 3B, 3C**; R^2^ = 0.72). No recombination was detected. Most of the new sequences (*n* = 28/31) clustered within a dominant clade of sequences from Rio de Janeiro state (named RJ1; *n* = 74, SH-aLRT = 88.4), while a few (*n* = 3/31) grouped in a smaller clade (RJ2; *n* = 4, SH-aLRT = 95.2) together with a publicly available sequence from Rio de Janeiro. Most sequences from Duque de Caxias clustered with sequences from other municipalities, such as Niterói and São João de Meriti, suggesting frequent viral spread within state borders.

**Figure 3:**
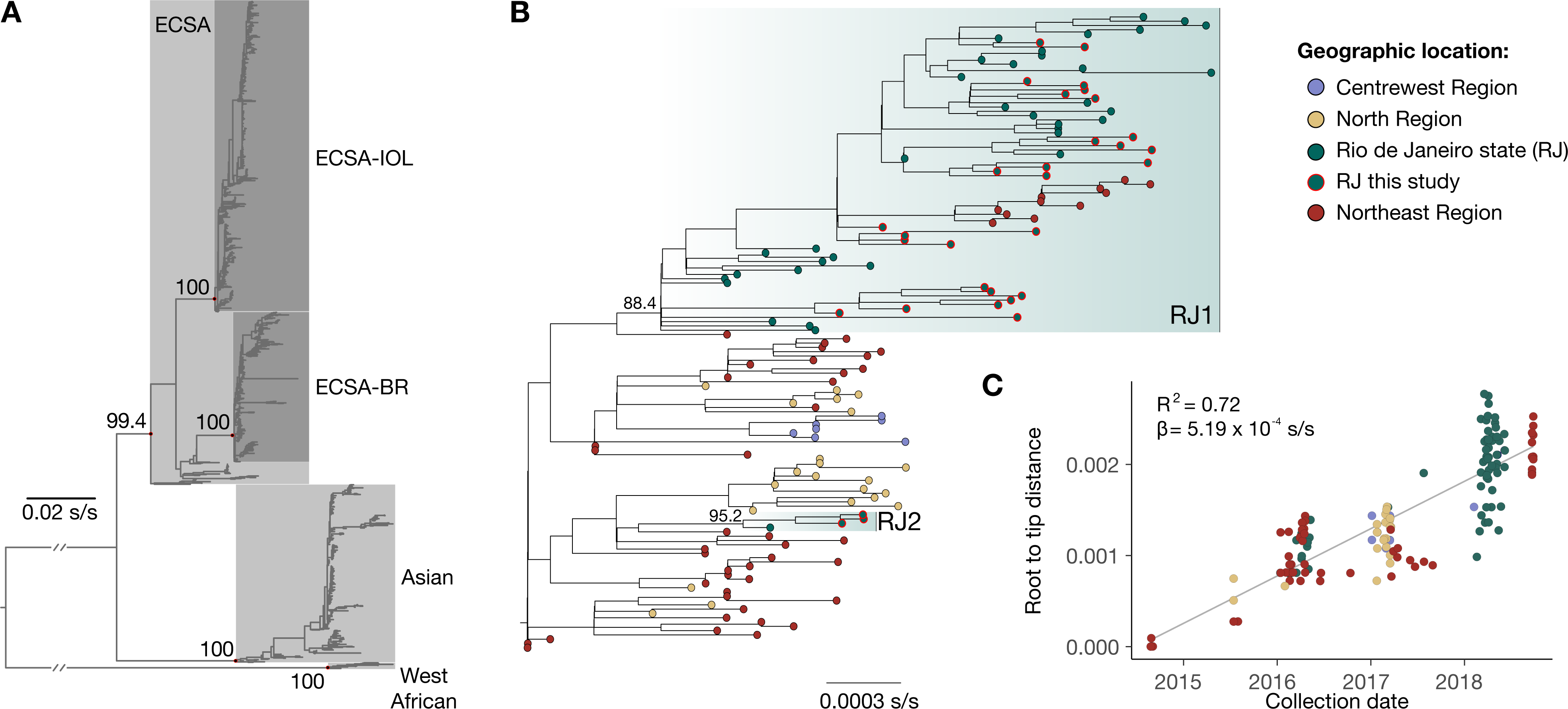
Maximum likelihood phylogenetic analysis of global and ECSA-American datasets. **(A)** Phylogenetic tree inferred from the global dataset. All new characterized genomes clustered within the ECSA-Br clade. Names of lineages and relevant clades are indicated. SH-aLRT statistical support values for these clades are shown close to their defining nodes (colored in red). **(B)** Phylogeny inferred from the filtered ECSA-American dataset. Tip shapes are colored according to sampling location (Centre-West region: purple, North region: yellow, Northeast region: red, Rio de Janeiro state: green). Sequences generated in this study are highlighted with red circles around the tip shapes. Clades composed mostly by RJ sequences are indicated along their SH-aLRT support values. **(C)** The root-to-tip regression plot, which indicates a strong temporal signal (R^2^ = 0.72, slope = 5.19 x 10^-4^). Scale bars represent substitutions per site (s/s).

Our time-scaled phylogenetic analysis estimated an evolutionary rate of around 5.72 x 10^-4^ (95% Bayesian credible interval, BCI, 4.89 x 10^-4^ – 6.50 x 10^-4^) substitutions per site per year and corroborated that the ECSA-American clade emerged in the Northeast region in mid-2014 (mean: 21 July, 95% BCI: 29 May to 25 August; **Figure 4A**). CHIKV-ECSA-American clade spread to the North and Centre-West regions of Brazil, as well as to Rio de Janeiro state. RJ1 and RJ2 clades result from independent introductions from the Northeast region with location posterior probability support > 99%. In contrast, the tMRCA for RJ1 was estimated in mid-2015 (mean: 7 July, 95% BCI: 4 April to 24 September 2015; clade posterior probability: 0.99), RJ2 tMRCA was dated around mid-2017 (mean: 7 July 2017, 95% BCI: 12 March 2017 to 6 November 2017; clade posterior probability: 1.00). Although we detected a more recent introduction from the Northeast region, our data suggests that the 2018 chikungunya epidemic in Rio de Janeiro was driven mostly by the RJ1 clade that was circulating in Rio de Janeiro since mid-2015.

**Figure 4:**
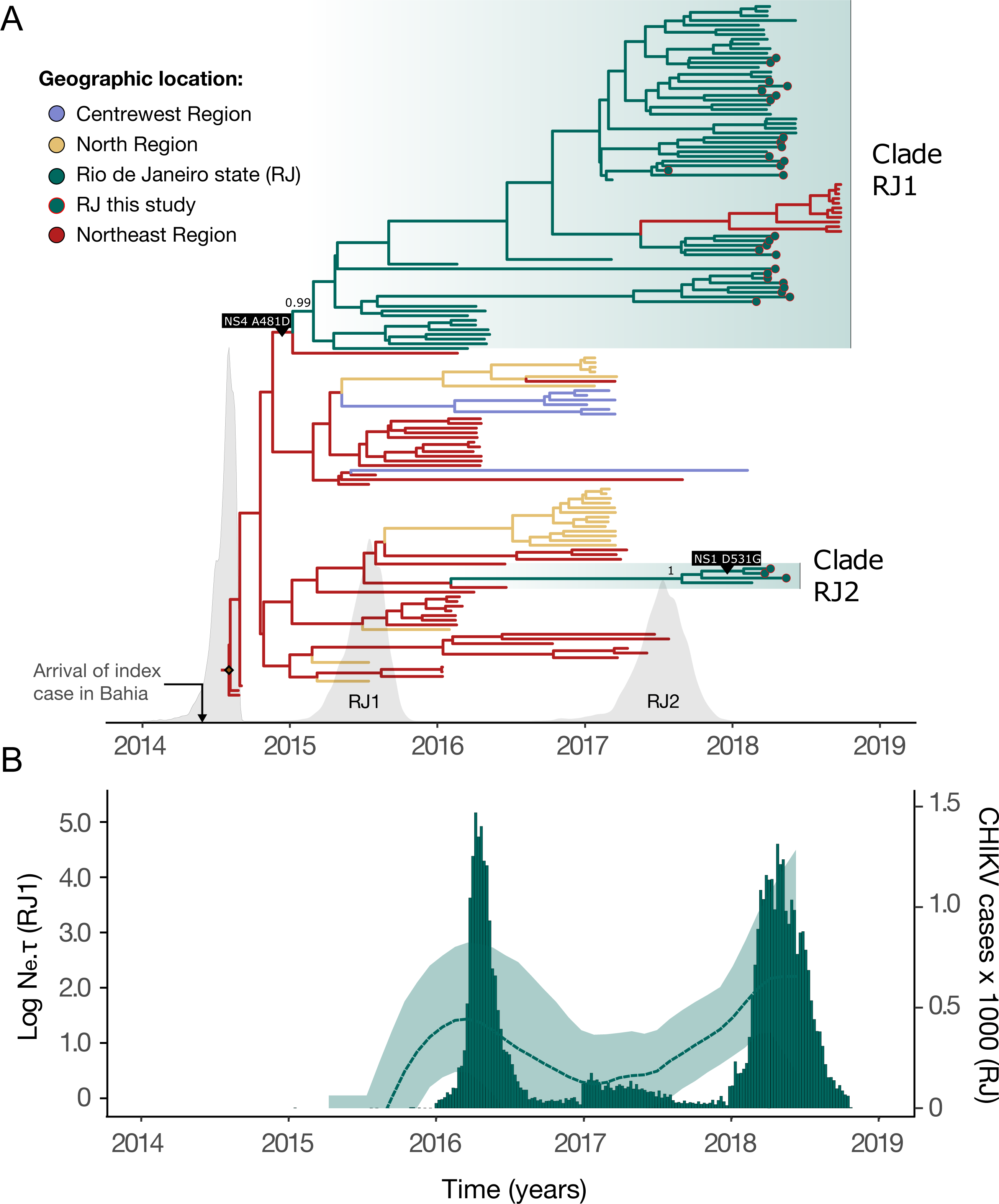
Bayesian time scaled phylogeographic reconstruction for the clade ECSA-Br. **(A)** The molecular clock phylogeny annotated with discrete trait reconstructions. Colors indicate estimated ancestral locations (Centre-West region: purple, North region: yellow, Northeast region: red, Rio de Janeiro state: green). Tip shapes mark sequences generated in this study. The x-axis depicts the timescale, while the density plots indicate the posterior distributions estimated for the age of clades ECSA-American, RJ1 and RJ2. Posterior probabilities for both RJ clades are shown. Positively selected mutations detected with MEME and FEL models (NS1: D351G and NS4: A481D) are exhibited on the branches where they occurred according to the ancestral states reconstruction performed (TreeTime). The inset marks the date of arrival of the index case in the Northeast region. **(B)** Skygrid reconstruction plot for the clade RJ1. A separate analysis was performed with only RJ sequences from the clade RJ1, allowing the reconstruction of the dynamics of variation of viral effective population size in the state (left y-axis). The analysis reveals CHIKV genetic diversity varied over time, with periods of high diversity matching peaks observed in incidence data (right y-axis).

The demographic pattern of the RJ1 clade inferred through coalescent non-parametric analysis, revealed that virus genetic diversity rose during the 2016 epidemic, declined through 2017, and rose again (**Figure 4B**). Notably, epidemiological and genetic data independently suggest that 2017 had a much lower number of infections than 2016 and 2018, despite the circulation of locally established lineage (RJ1) and a window of climatic suitability for transmission (**Figure 1D**).

### 3.4. Adaptive molecular evolution of CHIKV ECSA-American

Our ancestral reconstruction revealed 93 mutations associated with RJ lineages, of which 75 were synonymous and 18 non-synonymous (**Supplementary Figure S4**). Non-synonymous mutations have been identified in nearly all CHIKV ORFs: nsP1 (R76K, D531G), nsP2 (P16L, G261R, N299S, A545S, V645A, T652I), nsP3 (T422I, R435H), nsP4 (R99Q, I111V, S157C), C (K89R, A264T), E2 (V408I) and E1 (A305T, V669M). MEME and FEL models detected three and two amino acid changes under positive selection with statistical significance (*p* < 0.01; see also **Supplementary File S4**), respectively. Both models indicate that amino acids nsP1: D531G and nsP4: A481D were under positive selection. Interestingly, nsP4: A481D is a lineage-defining mutation of a clade that comprehends lineage RJ1 and a single sequence from Sergipe state (Northeast region, Accession Number: KY055011) (**Figure 4A, Supplementary Figure S4A**). In contrast, the nsP1: D531G amino acid change occurs within a subclade of RJ2, but also emerged multiple independent times within RJ1 (**Figure 4A, Supplementary Figure S4A**).

## 4. Discussion

We provide a detailed picture of the epidemic history of CHIKV ECSA-American in Rio de Janeiro state from 2016 to 2018, which could be divided into three epidemic phases. First, our analyses suggest that CHIKV ECSA was introduced in Rio de Janeiro around mid-2015, causing its first recorded epidemic in 2016, once ZIKV cases started decreasing in the state [64]. We estimated a *R_0_* for CHIKV ECSA-American in Rio de Janeiro to be around 1.56, similar to those reported to CHIKV across other settings [15,65–67]. Additionally, our R*_0_* for CHIKV was consistent with estimates for DENV in Rio de Janeiro [63], and lower compared to estimates for Zika [63], which caused large epidemics in Rio de Janeiro between 2015–2016 [68]. However, we then found unusually low CHIKV incidence and genetic diversity in 2017. The implementation of large-scale *Wolbachia* interventions in *Ae. aegypti* populations in Rio de Janeiro only started in late 2017 in a single neighborhood (*Ilha do Governador*); therefore it is unlikely to explain the observed patterns [69]. Low chikungunya incidences in 2017 were also observed in a large serosurveillance study of blood donors [70]. Thus, we hypothesize that the low number of cases in 2017 can partially be explained by climatic factors since *Ae. aegypti* suitability, mean temperature, or humidity in 2017 was lower compared to previous and following years. Moreover, the large chikungunya epidemic in Rio de Janeiro in 2018 indicates that the trend observed in 2017 was unlikely to be caused by population immunity. Indeed, surveys found that 18% of seroprevalence against CHIKV in >2,000 participants screened between July and October 2018 [71]. Additional investigations into the dynamics of arbovirus population immunity, including accurate estimates of force of infection, and analysis of data on vector control policies and population behavior, could help better understand the dynamics and evolution of CHIKV transmission in the Americas.

Overall, our results show that CHIKV incidence, R_t_, and *Ae. aegypti* transmission potential display a seasonal pattern and are generally synchronized. Similar trends have been noticed for ZIKV [15, 23], and further highlight the interplay between vector ecology, climate and arbovirus disease transmission. Given its major connectivity at the national and international level, Rio de Janeiro is a key hotspot for arbovirus transmission and a location of interest for epidemiological surveillance in Brazil. For instance, several events of dengue lineage replacement were caused by novel lineages initially detected in Rio de Janeiro [24]. In 2018, three different arboviruses circulated in Rio de Janeiro, CHIKV, DENV, and ZIKV, with CHIKV causing the most infections. We noticed a similar scenario in the Duque de Caxias municipality, from which 48.6% were positive for CHIKV and much smaller proportions for ZIKV (2.79%) and DENV (1.11%) in 2018. Overall, clinical symptoms in our cohort were similar to those described elsewhere [72]. However, among the CHIKV RNA-positive cases, we recorded one presenting neurological manifestations. A recent study has shown that neurological signs and symptoms were reported in 39% of CHIKV cases with a fatal outcome [3]. Additional studies are needed to investigate host and viral factors associated with clinical manifestations.

Our genetic analyses confirm that the clade ECSA-American lineage emerged in Northeast Brazil in mid-2014, consistent with the arrival of the index case in Bahia state in late May [13, 14]. This lineage then spread to all other regions over the following couple of years, being introduced in Rio de Janeiro around mid-2015, and later in mid-2017. We show that both introductions occurred from the Northeast region, consistent with previous findings [17, 18, 26]. As the first autochthonous cases of CHIKV in Rio de Janeiro were detected in November 2015, the period of cryptic transmission in the state was between two and seven months, consistent with previous estimates [17, 26].

Our analyses show the acquisition of a range of non-synonymous mutations in both non-structural and structural genes of the ECSA-American lineage. Although *Ae*. *albopictus* mosquitoes are abundant in Rio de Janeiro [73], none of the new sequences presented the mutations E1:A226V or E2:L210Q, previously associated with increased transmission competence by *Ae*. *albopictus* mosquitoes in the Indian Ocean Lineage [8, 9]. However, it remains unclear whether other mutations in CHIKV ECSA-American strains are associated with increased transmissibility in *Ae. aegypti* and/or *Ae. albopictus*. Notwithstanding, our analyses prove that at least two non-synonymous mutations associated with sequences from Brazil evolved through molecular adaptation: nsP1: D351G and nsP4: A481D. While evidence for positive selection in non-structural genes has already been identified for the CHIKV Asian genotype [74], this is the first report of this phenomenon for CHIKV ECSA. Signals of positive selection in non-structural proteins have also been detected for another alphavirus (Western equine encephalitis virus) in natural settings [75, 76], and experiments with chimeric alphaviruses have shown that adaptive mutations in nonstructural proteins are essential for viral replication and infectivity [77]. Nonstructural proteins have also been shown to be preferential targets for adaptive molecular evolution among different flaviviruses [78]. As these new mutations may impact the virus’ biological properties and epidemiology, further studies with cellular and animal models, including *Ae*. *aegypti* and *Ae*. *albopictus* vector competence surveys, should be performed to clarify their phenotypic impact.

In conclusion, our study sheds light on the epidemiological dynamics of chikungunya in the state of Rio de Janeiro up to 2018. We demonstrate that chikungunya incidence and transmissibility varied seasonally, following trends in *Ae*. *aegypti* transmission potential. We also provided first reports of CHIKV ECSA-American epidemiological parameters and identified mutations of interest that evolved under positive selection. We encourage scaling up diagnostic efforts for arboviruses and deployment of effective mosquito control strategies. Larger forthcoming genomic and epidemiological CHIKV-ECSA studies will help to disentangle the impact of virus molecular evolution and adaptation, host population behavior and immunity, and vector ecology to improve public health interventions against arboviruses.

## Data Availability

All data produced in the present work are contained in the manuscript

## 6 Acknowledgements

We thank the Health Department of the Municipality of Duque de Caxias, RJ, Brazil, which allowed the collection of clinical samples from patients with suspected arboviruses.

## 8 Author contributions

**Filipe Romero Rebello Moreira** (data curation, formal analysis, investigation, methodology, software, visualization, writing - original draft, writing - review and editing), **Mariane Talon de Menezes** (data curation, investigation, methodology, writing - review and editing), **Clarisse Salgado-Benvindo** (data curation, investigation, methodology, writing - review and editing), **Charles Whittaker** (formal analysis, investigation writing - review and editing), **Victoria Cox** (formal analysis, investigation writing - review and editing), **Nilani Chandradeva** (formal analysis, investigation, writing - review and editing), **Hury Hellen Souza de Paula** (data curation, investigation, writing - review and editing), **André Frederico Martins** (data curation, investigation, writing - review and editing), **Raphael Rangel das Chagas** (data curation, investigation, writing - review and editing), **Rodrigo Decembrino Vargas Brasil** (data curation, investigation, writing - review and editing), **Darlan Silva Cândido** (formal analysis, methodology, software, writing - review and editing), **Alice Laschuk Herlinger** (investigation, writing - review and editing), **Marisa de Oliveira Ribeiro** (methodology, writing - review and editing), **Monica Barcellos Arruda** (methodology, writing - review and editing), **Patricia Alvarez** (methodology, writing - review and editing), **Marcelo Calado de Paula** (investigation, writing - review and editing), **Ilaria Dorigatti** (formal analysis, investigation, writing - review and editing), **Oliver Brady** (formal analysis, investigation, writing - review and editing), **Carolina Moreira Voloch** (formal analysis, methodology, resources, writing - review and editing), **Amilcar Tanuri** (funding acquisition, resources, writing - review and editing), **Felipe Iani** (data curation, writing - review and editing), **William Marciel de Souza** (investigation, writing - review and editing), **Sergian Vianna Cardozo** (conceptualization, data curation, investigation, methodology, project administration, supervision, writing - review and editing), **Nuno Rodrigues Faria** (conceptualization, formal analysis, methodology, resources, supervision, visualization, writing - review and editing), **Renato Santana de Aguiar** (conceptualization, funding acquisition, methodology, project administration, resources, supervision, writing - review and editing).

---

**Supplementary Figure S1:** CHIKV incidence within Rio de Janeiro state. **(A)** CHIKV incidence per state region. Rio de Janeiro is officially divided into five intermediate regions, each comprehending between 12 and 26 municipalities (IBGE, https://www.ibge.gov.br/apps/regioes_geograficas/, last accessed 17 August 2022). **(B)** CHIKV incidence per municipality. As the state comprehends 92 municipalities, and many of them present low incidence levels through the entire period, individual color legends have been omitted. Municipalities that at any point presented weekly incidence above 500 cases were highlighted in the plot (Campos dos Goytacazes, Rio de Janeiro, São Gonçalo).

**Supplementary Figure S2:** *R_0_* estimates and sensitivity analysis. *R_0_* estimates obtained with epidemiological data under different generation time distributions (gamma distributions with means 10, 14 and 20, and constant standard deviation: 6.4 days).

**Supplementary Figure S3:** *R_t_* estimates and sensitivity analysis. **(A)** *R_t_* estimate performed with different generation time (GT) distributions (gamma distributions with means 10, 14 and 20, and constant standard deviation: 6.4 days). Colors indicate estimates for different GTs (10: gray, 14: yellow, 20: blue). **(B)** *R_t_* estimates performed with different sliding window lengths (3 to 8 weeks). Colors indicate estimates for different window lengths (3: salmon, 4: yellow, 5: green, 6: blue, 7: purple, 8: pink). Solid lines indicate mean values and ribbons indicate the 95% confidence intervals.The dashed lines denote the critical epidemic threshold (*R_t_* = 1).

**Supplementary Figure S4:** TreeTime and HyPhy analysis. **(A)** The molecular clock tree had its branches colored according to the ancestral location’s reconstructions. Tip shapes indicate sequences generated in this study. Light gray ticks along branches indicate synonymous mutations, while dark gray ticks mark non-synonymous mutations. Mutations for which evidence of positive selection was found (MEME and FEL models) are marked in red (NS4: A481D) and blue (NS1: D531G). **(B)** CHIKV genome scheme exhibiting all mutations reconstructed and associated with RJ clades. Non-synonymous mutations are numbered from 1 to 18 and their positions and corresponding amino acid replacements are indicated by black vertical lines. Similarly, vertical gray lines indicate the position of the synonymous mutations inferred. Non-structural and structural open reading frames are colored in dark green and light green, respectively.

**Supplementary File S1:** Data and R code used in epidemiological analyses.

**Supplementary File S2:** BEAST xml files and outputs generated in this study.

**Supplementary File S3:** Clinical data from CHIKV positive patients.

**Supplementary File S4:** Input and output files of selection analysis with Datamonkey.

## 5. References

1. Pialoux G, Gaüzère B-A, Jauréguiberry S, Strobel M. Chikungunya, an epidemic arbovirosis. Lancet Infect Dis. 2007;7: 319–327. doi:10.1016/S1473-3099(07)70107-X

2. Chen R, Mukhopadhyay S, Merits A, Bolling B, Nasar F, Coffey LL, et al. ICTV Virus Taxonomy Profile: Togaviridae. J Gen Virol. 2018;99: 761–762. doi:10.1099/jgv.0.001072

3. de Lima STS, de Souza WM, Cavalcante JW, da Silva Candido D, Fumagalli MJ, Carrera J-P, et al. Fatal Outcome of Chikungunya Virus Infection in Brazil. Clin Infect Dis. 2021;73: e2436–e2443. doi:10.1093/cid/ciaa1038

4. Nsoesie EO, Kraemer MU, Golding N, Pigott DM, Brady OJ, Moyes CL, et al. Global distribution and environmental suitability for chikungunya virus, 1952 to 2015. Euro Surveill. 2016;21. doi:10.2807/1560-7917.ES.2016.21.20.30234

5. Grubaugh ND, Faria NR, Andersen KG, Pybus OG. Genomic Insights into Zika Virus Emergence and Spread. Cell. 2018;172: 1160–1162. doi:10.1016/j.cell.2018.02.027

6. Volk SM, Chen R, Tsetsarkin KA, Adams AP, Garcia TI, Sall AA, et al. Genome-scale phylogenetic analyses of chikungunya virus reveal independent emergences of recent epidemics and various evolutionary rates. J Virol. 2010;84: 6497–6504. doi:10.1128/JVI.01603-09

7. Pybus OG, Tatem AJ, Lemey P. Virus evolution and transmission in an ever more connected world. Proc Biol Sci. 2015;282: 20142878. doi:10.1098/rspb.2014.2878

8. Tsetsarkin KA, Vanlandingham DL, McGee CE, Higgs S. A single mutation in chikungunya virus affects vector specificity and epidemic potential. PLoS Pathog. 2007;3: e201. doi:10.1371/journal.ppat.0030201

9. Tsetsarkin KA, McGee CE, Volk SM, Vanlandingham DL, Weaver SC, Higgs S. Epistatic roles of E2 glycoprotein mutations in adaption of chikungunya virus to Aedes albopictus and Ae. aegypti mosquitoes. PLoS One. 2009;4: e6835. doi:10.1371/journal.pone.0006835

10. Tsetsarkin KA, Chen R, Leal G, Forrester N, Higgs S, Huang J, et al. Chikungunya virus emergence is constrained in Asia by lineage-specific adaptive landscapes. Proc Natl Acad Sci U S A. 2011;108: 7872–7877. doi:10.1073/pnas.1018344108

11. Leparc-Goffart I, Nougairede A, Cassadou S, Prat C, de Lamballerie X. Chikungunya in the Americas. The Lancet. 2014. p. 514. doi:10.1016/S0140-6736(14)60185-9

12. Pan American Health Organization (PAHO). Chikungunya. [cited 14 Aug 2022]. Available: https://www.paho.org/en/topics/chikungunya

13. Nunes MRT, Faria NR, de Vasconcelos JM, Golding N, Kraemer MUG, de Oliveira LF, et al. Emergence and potential for spread of Chikungunya virus in Brazil. BMC Med. 2015;13: 102. doi:10.1186/s12916-015-0348-x

14. Faria NR, Lourenço J, de Cerqueira EM, de Lima MM, Pybus O, Alcantara LCJ. Epidemiology of Chikungunya Virus in Bahia, Brazil, 2014-2015Vaccine Hesitancy CollectionPLOS Science Reddit AMAHealthMap EbolaNew Twitter. PLoS Curr. 2016. doi:10.1371/currents.outbreaks.c97507e3e48efb946401755d468c28b2

15. Naveca FG, Claro I, Giovanetti M, de Jesus JG, Xavier J, Iani FC de M, et al. Genomic, epidemiological and digital surveillance of Chikungunya virus in the Brazilian Amazon. PLoS Negl Trop Dis. 2019;13: e0007065. doi:10.1371/journal.pntd.0007065

16. Charlys da Costa A, Thézé J, Komninakis SCV, Sanz-Duro RL, Felinto MRL, Moura LCC, et al. Spread of Chikungunya Virus East/Central/South African Genotype in Northeast Brazil. Emerg Infect Dis. 2017;23: 1742–1744. doi:10.3201/eid2310.170307

17. Xavier J, Giovanetti M, Fonseca V, Thézé J, Gräf T, Fabri A, et al. Circulation of chikungunya virus East/Central/South African lineage in Rio de Janeiro, Brazil. PLOS ONE. 2019. p. e0217871. doi:10.1371/journal.pone.0217871

18. Souza TML, Vieira YR, Delatorre E, Barbosa-Lima G, Luiz RLF, Vizzoni A, et al. Emergence of the East-Central-South-African genotype of Chikungunya virus in Brazil and the city of Rio de Janeiro may have occurred years before surveillance detection. Sci Rep. 2019;9: 2760. doi:10.1038/s41598-019-39406-9

19. Vasconcellos AF, Silva JMF, de Oliveira AS, Prado PS, Nagata T, Resende RO. Genome sequences of chikungunya virus isolates circulating in midwestern Brazil. Arch Virol. 2019;164: 1205–1208. doi:10.1007/s00705-019-04174-4

20. de Souza WM, de Lima STS, Simões Mello LM, Candido DS, Buss L, Whittaker C, et al. Spatiotemporal dynamics and recurrence of chikungunya virus in Brazil: an epidemiological study. Lancet Microbe. 2023. doi:10.1016/S2666-5247(23)00033-2

21. Gräf T, Vazquez C, Giovanetti M, de Bruycker-Nogueira F, Fonseca V, Claro IM, et al. Epidemiologic History and Genetic Diversity Origins of Chikungunya and Dengue Viruses, Paraguay. Emerg Infect Dis. 2021;27: 1393–1404. doi:10.3201/eid2705.204244

22. White SK, Mavian C, Salemi M, Morris JG Jr, Elbadry MA, Okech BA, et al. A new “American” subgroup of African-lineage Chikungunya virus detected in and isolated from mosquitoes collected in Haiti, 2016. PLoS One. 2018;13: e0196857. doi:10.1371/journal.pone.0196857

23. Faria NR, Quick J, Claro IM, Thézé J, de Jesus JG, Giovanetti M, et al. Establishment and cryptic transmission of Zika virus in Brazil and the Americas. Nature. 2017;546: 406–410. doi:10.1038/nature22401

24. Torres MC, de Bruycker Nogueira F, Fernandes CA, Louzada Silva Meira G, Ferreira de Aguiar S, Chieppe AO, et al. Re-introduction of dengue virus serotype 2 in the state of Rio de Janeiro after almost a decade of epidemiological silence. PLoS One. 2019;14: e0225879. doi:10.1371/journal.pone.0225879

25. Brasil, 2019. Boletim Epidemiológico—Monitoramento dos casos de dengue, febre de chikungunya e doença aguda pelo vírus Zika até a Semana Epidemiológica 52 de 2018. Ministério da Saúde. Secretaria de Vigilância em Saúde, report no.: 4.

26. Fabri AA, Rodrigues CDDS, Santos CCD, Chalhoub FLL, Sampaio SA, Faria NR da C, et al. Co-Circulation of Two Independent Clades and Persistence of CHIKV-ECSA Genotype during Epidemic Waves in Rio de Janeiro, Southeast Brazil. Pathogens. 2020;9. doi:10.3390/pathogens9120984

27. Wallinga J, Lipsitch M. How generation intervals shape the relationship between growth rates and reproductive numbers. Proc Biol Sci. 2007;274: 599–604. doi:10.1098/rspb.2006.3754

28. Cori A, Ferguson NM, Fraser C, Cauchemez S. A new framework and software to estimate time-varying reproduction numbers during epidemics. Am J Epidemiol. 2013;178: 1505–1512. doi:10.1093/aje/kwt133

29. Salje H, Lessler J, Paul KK, Azman AS, Rahman MW, Rahman M, et al. How social structures, space, and behaviors shape the spread of infectious diseases using chikungunya as a case study. Proc Natl Acad Sci U S A. 2016;113: 13420–13425. doi:10.1073/pnas.1611391113

30. R Core Team (2022). R: A language and environment for statistical computing. R Foundation for Statistical Computing, Vienna, Austria. https://www.R-project.org/.

31. Obadia T, Haneef R, Boëlle P-Y. The R0 package: a toolbox to estimate reproduction numbers for epidemic outbreaks. BMC Med Inform Decis Mak. 2012;12: 147. doi:10.1186/1472-6947-12-147

32. Copernicus climate data store. [cited 23 Feb 2023]. Available: https://cds.climate.copernicus.eu/cdsapp#!/dataset/reanalysis-era5-single-levels?tab=overview

33. Alduchov OA, Eskridge RE. Improved Magnus Form Approximation of Saturation Vapor Pressure. J Appl Meteorol Climatol. 1996;35: 601–609. doi:10.1175/1520-0450(1996)035<0601:IMFAOS>2.0.CO;2

34. August, E. F. Ueber die Berechnung der Expansivkraft des Wasserdunstes. Ann. Phys. Chem. 1828;13: 122–137.

35. Magnus, G. Versuche über die Spannkräfte des Wasserdampfs. Ann. Phys. Chem. 1844;61: 225–247.

36. Obolski U, Perez PN, Villabona-Arenas CJ, Thézé J, Faria NR, Lourenço J. MVSE: An R-package that estimates a climate-driven mosquito-borne viral suitability index. Methods Ecol Evol. 2019;10: 1357–1370. doi:10.1111/2041-210X.13205

37. Quick J, Grubaugh ND, Pullan ST, Claro IM, Smith AD, Gangavarapu K, et al. Multiplex PCR method for MinION and Illumina sequencing of Zika and other virus genomes directly from clinical samples. Nat Protoc. 2017;12: 1261–1276. doi:10.1038/nprot.2017.066

38. Bolger AM, Lohse M, Usadel B. Trimmomatic: a flexible trimmer for Illumina sequence data. Bioinformatics. 2014;30: 2114–2120. doi:10.1093/bioinformatics/btu170

39. Langmead B, Salzberg SL. Fast gapped-read alignment with Bowtie 2. Nat Methods. 2012;9: 357–359. doi:10.1038/nmeth.1923

40. Li H, Handsaker B, Wysoker A, Fennell T, Ruan J, Homer N, et al. The Sequence Alignment/Map format and SAMtools. Bioinformatics. 2009;25: 2078–2079. doi:10.1093/bioinformatics/btp352

41. Li H. A statistical framework for SNP calling, mutation discovery, association mapping and population genetical parameter estimation from sequencing data. Bioinformatics. 2011;27: 2987–2993. doi:10.1093/bioinformatics/btr509

42. Quinlan AR, Hall IM. BEDTools: a flexible suite of utilities for comparing genomic features. Bioinformatics. 2010;26: 841–842. doi:10.1093/bioinformatics/btq033

43. Sayers EW, Bolton EE, Brister JR, Canese K, Chan J, Comeau DC, et al. Database resources of the national center for biotechnology information. Nucleic Acids Res. 2022;50: D20–D26. doi:10.1093/nar/gkab1112

44. Katoh K, Standley DM. MAFFT multiple sequence alignment software version 7: improvements in performance and usability. Mol Biol Evol. 2013;30: 772–780. doi:10.1093/molbev/mst010

45. Minh BQ, Schmidt HA, Chernomor O, Schrempf D, Woodhams MD, von Haeseler A, et al. IQ-TREE 2: New Models and Efficient Methods for Phylogenetic Inference in the Genomic Era. Mol Biol Evol. 2020;37: 1530–1534. doi:10.1093/molbev/msaa015

46. Kalyaanamoorthy S, Minh BQ, Wong TKF, von Haeseler A, Jermiin LS. ModelFinder: fast model selection for accurate phylogenetic estimates. Nat Methods. 2017;14: 587–589. doi:10.1038/nmeth.4285

47. Guindon S, Dufayard J-F, Lefort V, Anisimova M, Hordijk W, Gascuel O. New algorithms and methods to estimate maximum-likelihood phylogenies: assessing the performance of PhyML 3.0. Syst Biol. 2010;59: 307–321. doi:10.1093/sysbio/syq010

48. Martin DP, Murrell B, Golden M, Khoosal A, Muhire B. RDP4: Detection and analysis of recombination patterns in virus genomes. Virus Evol. 2015;1: vev003. doi:10.1093/ve/vev003

49. Rambaut A, Lam TT, Max Carvalho L, Pybus OG. Exploring the temporal structure of heterochronous sequences using TempEst (formerly Path-O-Gen). Virus Evol. 2016;2: vew007. doi:10.1093/ve/vew007

50. Suchard MA, Lemey P, Baele G, Ayres DL, Drummond AJ, Rambaut A. Bayesian phylogenetic and phylodynamic data integration using BEAST 1.10. Virus Evol. 2018;4: vey016. doi:10.1093/ve/vey016

51. Lemey P, Rambaut A, Drummond AJ, Suchard MA. Bayesian phylogeography finds its roots. PLoS Comput Biol. 2009;5: e1000520. doi:10.1371/journal.pcbi.1000520

52. Hasegawa M, Yano T-A, Kishino H. A New Molecular Clock of Mitochondrial DNA and the Evolution of Hominoids. Proc Jpn Acad Ser B Phys Biol Sci. 1984;60: 95–98. doi:10.2183/pjab.60.95

53. Yang Z. Maximum likelihood phylogenetic estimation from DNA sequences with variable rates over sites: approximate methods. J Mol Evol. 1994;39: 306–314. doi:10.1007/BF00160154

54. Gill MS, Lemey P, Faria NR, Rambaut A, Shapiro B, Suchard MA. Improving Bayesian population dynamics inference: a coalescent-based model for multiple loci. Mol Biol Evol. 2013;30: 713–724. doi:10.1093/molbev/mss265

55. Ayres DL, Darling A, Zwickl DJ, Beerli P, Holder MT, Lewis PO, et al. BEAGLE: an application programming interface and high-performance computing library for statistical phylogenetics. Syst Biol. 2012;61: 170–173. doi:10.1093/sysbio/syr100

56. Rambaut A, Drummond AJ, Xie D, Baele G, Suchard MA. Posterior Summarization in Bayesian Phylogenetics Using Tracer 1.7. Syst Biol. 2018;67: 901–904. doi:10.1093/sysbio/syy032

57. Candido DS, Claro IM, de Jesus JG, Souza WM, Moreira FRR, Dellicour S, et al. Evolution and epidemic spread of SARS-CoV-2 in Brazil. Science. 2020;369: 1255–1260. doi:10.1126/science.abd2161

58. Murrell B, Wertheim JO, Moola S, Weighill T, Scheffler K, Kosakovsky Pond SL. Detecting individual sites subject to episodic diversifying selection. PLoS Genet. 2012;8: e1002764. doi:10.1371/journal.pgen.1002764

59. Kosakovsky Pond SL, Frost SDW. Not so different after all: a comparison of methods for detecting amino acid sites under selection. Mol Biol Evol. 2005;22: 1208–1222. doi:10.1093/molbev/msi105

60. Weaver S, Shank SD, Spielman SJ, Li M, Muse SV, Kosakovsky Pond SL. Datamonkey 2.0: A Modern Web Application for Characterizing Selective and Other Evolutionary Processes. Mol Biol Evol. 2018;35: 773–777. doi:10.1093/molbev/msx335

61. Kosakovsky Pond SL, Poon AFY, Velazquez R, Weaver S, Hepler NL, Murrell B, et al. HyPhy 2.5-A Customizable Platform for Evolutionary Hypothesis Testing Using Phylogenies. Mol Biol Evol. 2020;37: 295–299. doi:10.1093/molbev/msz197

62. Sagulenko P, Puller V, Neher RA. TreeTime: Maximum-likelihood phylodynamic analysis. Virus Evol. 2018;4: vex042. doi:10.1093/ve/vex042

63. Villela DAM, Bastos LS, DE Carvalho LM, Cruz OG, Gomes MFC, Durovni B, et al. Zika in Rio de Janeiro: Assessment of basic reproduction number and comparison with dengue outbreaks. Epidemiol Infect. 2017;145: 1649–1657. doi:10.1017/S0950268817000358

64. Freitas LP, Cruz OG, Lowe R, Sá Carvalho M. Space-time dynamics of a triple epidemic: dengue, chikungunya and Zika clusters in the city of Rio de Janeiro. Proc Biol Sci. 2019;286: 20191867. doi:10.1098/rspb.2019.1867

65. Salje H, Cauchemez S, Alera MT, Rodriguez-Barraquer I, Thaisomboonsuk B, Srikiatkhachorn A, et al. Reconstruction of 60 Years of Chikungunya Epidemiology in the Philippines Demonstrates Episodic and Focal Transmission. J Infect Dis. 2016;213: 604–610. doi:10.1093/infdis/jiv470

66. Perkins TA, Alex Perkins T, Metcalf CJE, Grenfell BT, Tatem AJ. Estimating Drivers of Autochthonous Transmission of Chikungunya Virus in its Invasion of the Americas. PLoS Currents. 2015. doi:10.1371/currents.outbreaks.a4c7b6ac10e0420b1788c9767946d1fc

67. Robinson M, Conan A, Duong V, Ly S, Ngan C, Buchy P, et al. A model for a chikungunya outbreak in a rural Cambodian setting: implications for disease control in uninfected areas. PLoS Negl Trop Dis. 2014;8: e3120. doi:10.1371/journal.pntd.0003120

68. de Oliveira WK, Carmo EH, Henriques CM, Coelho G, Vazquez E, Cortez-Escalante J, et al. Zika Virus Infection and Associated Neurologic Disorders in Brazil. N Engl J Med. 2017;376: 1591–1593. doi:10.1056/NEJMc1608612

69. Ribeiro Dos Santos G, Durovni B, Saraceni V, Souza Riback TI, Pinto SB, Anders KL, et al. Estimating the effect of the wMel release programme on the incidence of dengue and chikungunya in Rio de Janeiro, Brazil: a spatiotemporal modelling study. Lancet Infect Dis. 2022;22: 1587–1595. doi:10.1016/S1473-3099(22)00436-4

70. Custer B, Grebe E, Buccheri R, Bakkour S, Stone M, Capuani L, et al. Surveillance for Zika, chikungunya and dengue virus incidence and RNAemia in blood donors at four Brazilian blood centers during 2016-2019. J Infect Dis. 2022. doi:10.1093/infdis/jiac173

71. Périssé ARS, Souza-Santos R, Duarte R, Santos F, de Andrade CR, Rodrigues NCP, et al. Zika, dengue and chikungunya population prevalence in Rio de Janeiro city, Brazil, and the importance of seroprevalence studies to estimate the real number of infected individuals. PLoS One. 2020;15: e0243239. doi:10.1371/journal.pone.0243239

72. Mascarenhas M, Garasia S, Berthiaume P, Corrin T, Greig J, Ng V, et al. A scoping review of published literature on chikungunya virus. PLoS One. 2018;13: e0207554. doi:10.1371/journal.pone.0207554

73. Kraemer MUG, Reiner RC Jr, Brady OJ, Messina JP, Gilbert M, Pigott DM, et al. Past and future spread of the arbovirus vectors Aedes aegypti and Aedes albopictus. Nat Microbiol. 2019;4: 854–863. doi:10.1038/s41564-019-0376-y

74. Sahadeo NSD, Allicock OM, De Salazar PM, Auguste AJ, Widen S, Olowokure B, et al. Understanding the evolution and spread of chikungunya virus in the Americas using complete genome sequences. Virus Evol. 2017;3: vex010. doi:10.1093/ve/vex010

75. Bergren NA, Auguste AJ, Forrester NL, Negi SS, Braun WA, Weaver SC. Western equine encephalitis virus: evolutionary analysis of a declining alphavirus based on complete genome sequences. J Virol. 2014;88: 9260–9267. doi:10.1128/JVI.01463-14

76. Bergren NA, Haller S, Rossi SL, Seymour RL, Huang J, Miller AL, et al. “Submergence” of Western equine encephalitis virus: Evidence of positive selection argues against genetic drift and fitness reductions. PLoS Pathog. 2020;16: e1008102. doi:10.1371/journal.ppat.1008102

77. Teppor M, Žusinaite E, Karo-Astover L, Omler A, Rausalu K, Lulla V, et al. Semliki Forest Virus Chimeras with Functional Replicase Modules from Related Alphaviruses Survive by Adaptive Mutations in Functionally Important Hot Spots. J Virol. 2021;95: e0097321. doi:10.1128/JVI.00973-21

78. Sironi M, Forni D, Clerici M, Cagliani R. Nonstructural Proteins Are Preferential Positive Selection Targets in Zika Virus and Related Flaviviruses. PLoS Negl Trop Dis. 2016;10: e0004978. doi:10.1371/journal.pntd.0004978

