## Supplementary figures and images for "Epidemiological and genomic investigation of chikungunya virus in Rio de Janeiro state, Brazil, between 2015 and 2018"

### Supplementary Figure S1

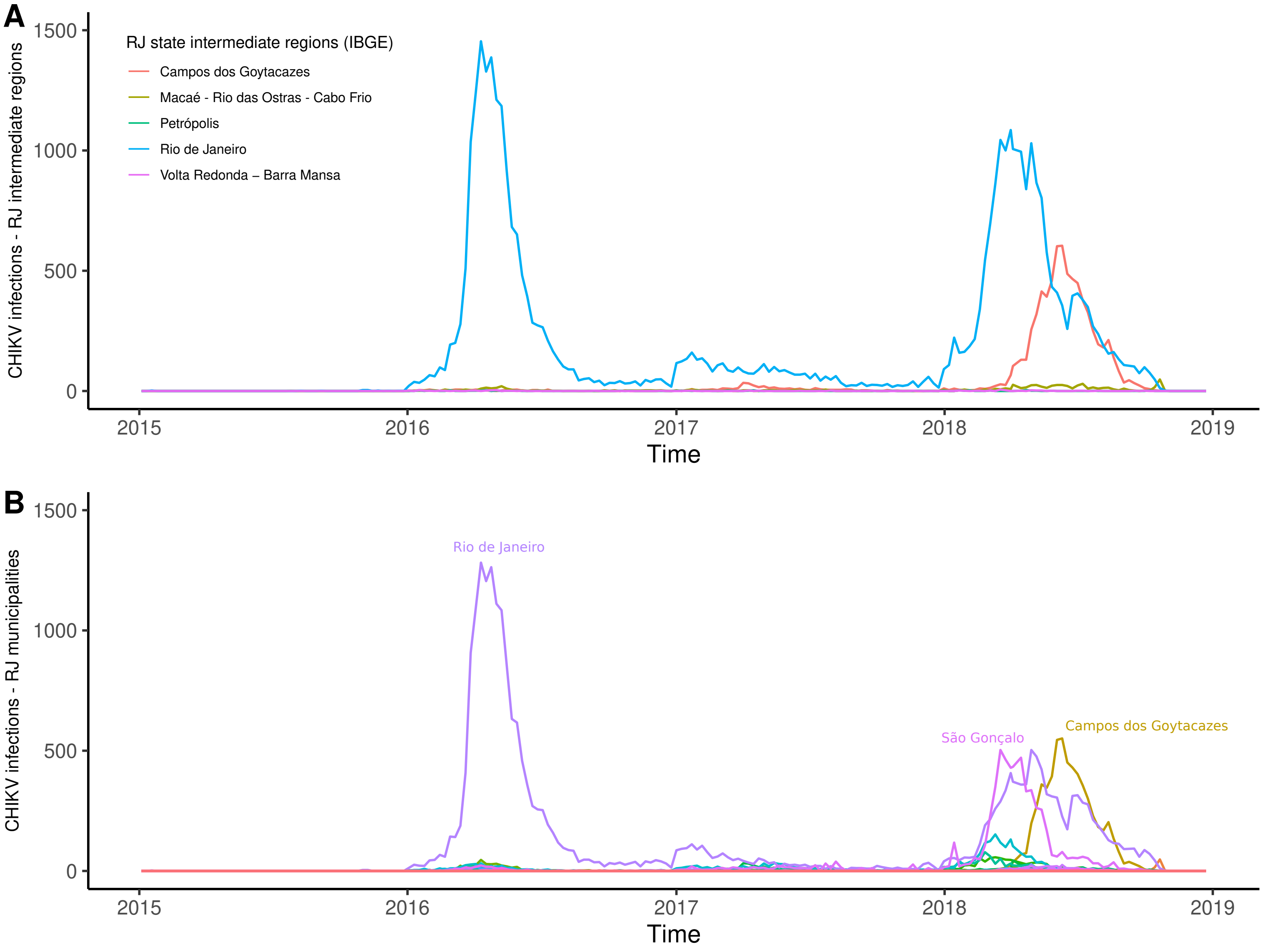

### Supplementary Figure S2

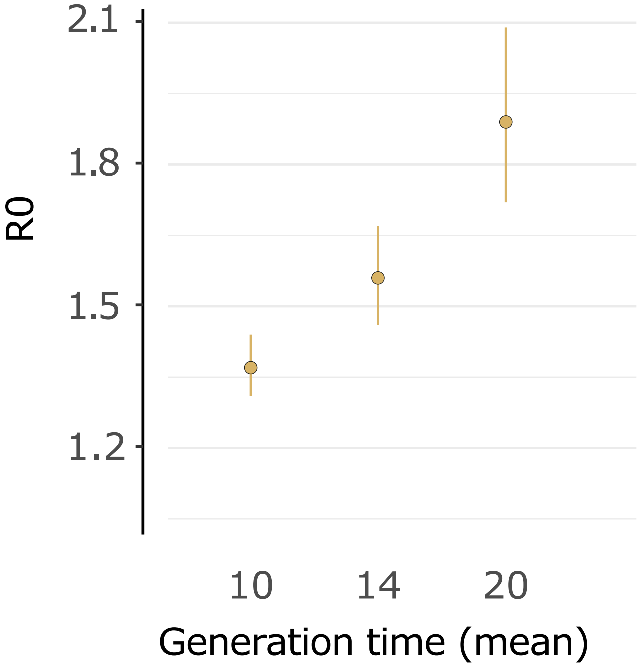

### Supplementary Figure S3

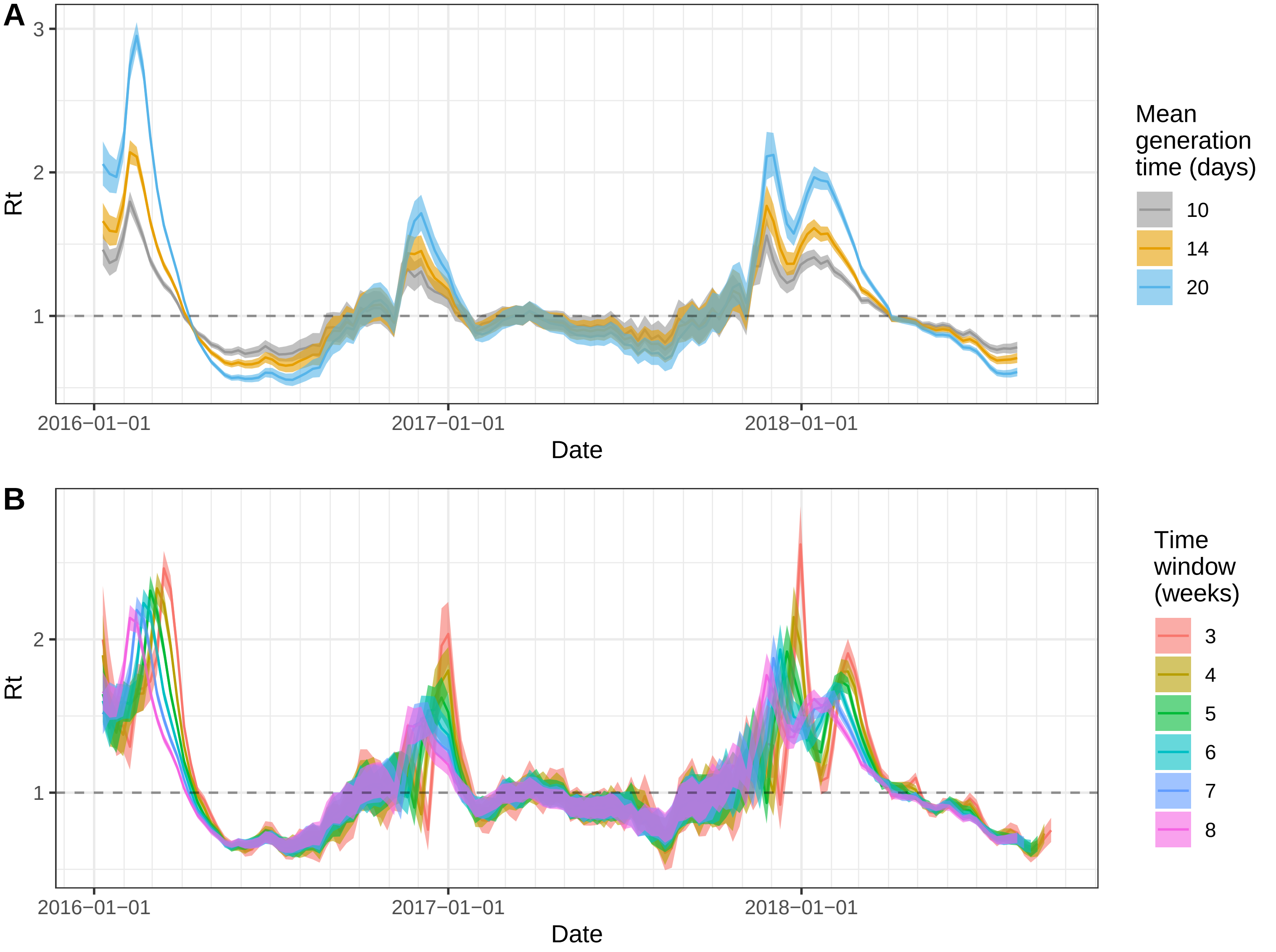

### Supplementary Figure S4

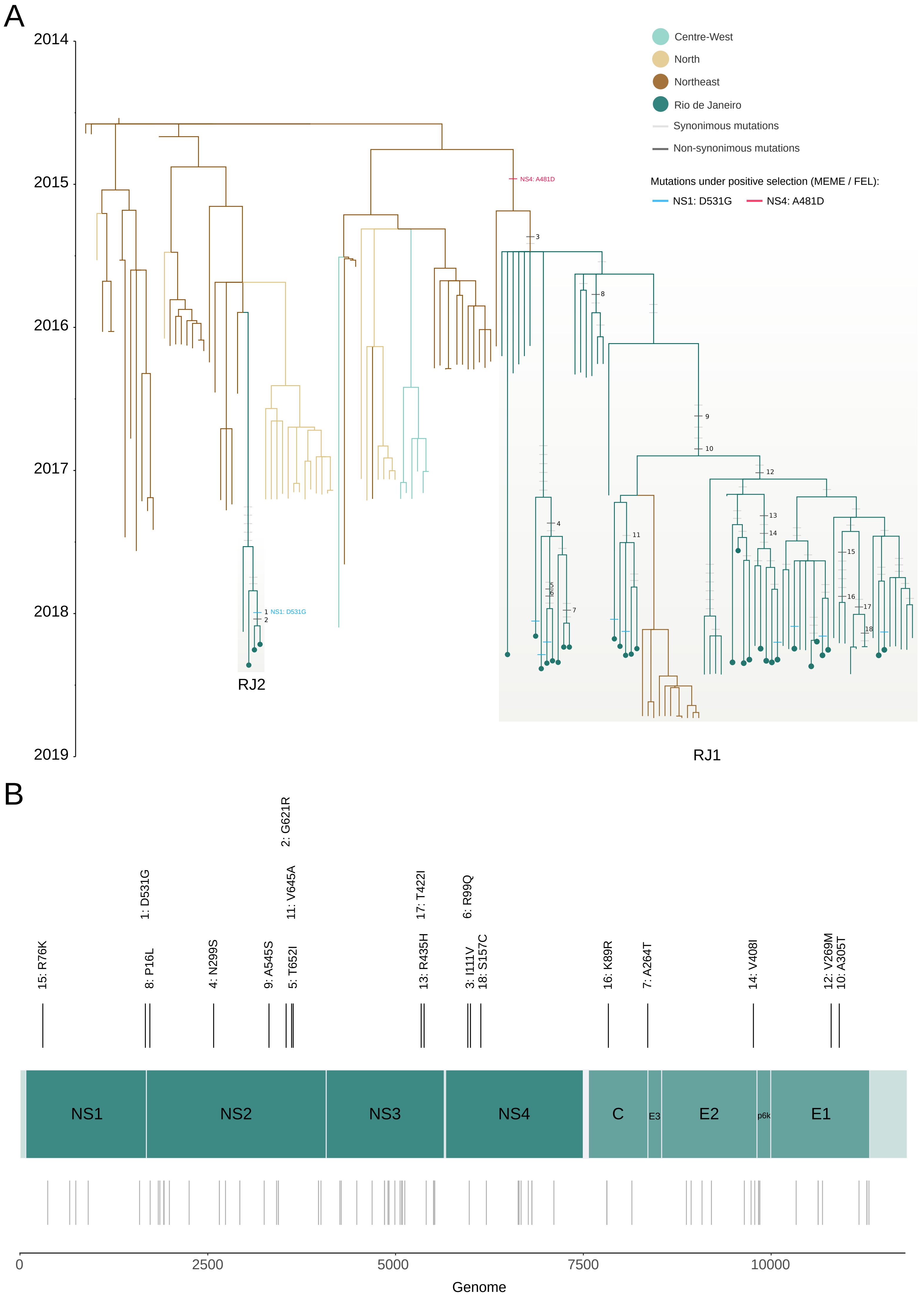
