## Supplementary File S1 for "Epidemiological and genomic investigation of chikungunya virus in Rio de Janeiro state, Brazil, between 2015 and 2018": Estimate_index_p.R

```
# Estimating indexP for RJ state - Brazil

source("MVSE.R")
require("pbapply")
require("scales")
require("genlasso")
require(tidyverse)
require(lubridate)

#analysis for RJ####
dat_RJ <- read.csv("data/RJ_state_weather_data_daily_2014_2018.csv", header = TRUE)
dat_RJ_clean <- dat_RJ %>%
  select(Date, d2m, t2m, RH) %>%
  rename(daily_min_temp = d2m, daily_max_temp = t2m, H = RH) %>% #MVSE requires these column names
  mutate(R = 0)#need this R column for MVSE to run (latest package updates require a rainfall input for other MVSE outputs)  

#calc daily mean temp using daily min and max
dat_RJ_cleaner <- dat_RJ_clean %>%
  rowwise() %>% #take row means of daily min and max temp to get mean daily temperature
  mutate(T = mean(c(daily_min_temp, daily_max_temp))) %>%  #we call this mean daily temperature, T
  separate(Date, c("year", "month", "day"), remove = FALSE) %>% #get day month year in sep columns 
  rename(date = Date)
dat_RJ_input <- data.frame(dat_RJ_cleaner$T, dat_RJ_cleaner$H, 
                           dat_RJ_cleaner$year, dat_RJ_cleaner$month,
                           dat_RJ_cleaner$day,
                           dat_RJ_cleaner$date, dat_RJ_cleaner$R)
names(dat_RJ_input) <- c("T", "H", "year", "month", "day", "date", "R")

#write this out as a CSV
write.csv(dat_RJ_input, file = "RJ/RJ_clim_clean.csv", row.names = FALSE) #this is the cleaned climate data that is fed into MVSE

#indexP analysis RJ####
detach("package:dplyr", unload = TRUE) #conflicts between MVSE and dplyr
setEmpiricalClimateSeries(filepath = 'RJ/RJ_clim_clean.csv')
setOutputFilePathAndTag('RJ/')

#ento parameters (leave as package defaults)
setMosqLifeExpPrior(pmean=14, psd=3, pdist='gaussian')  
setMosqIncPerPrior(pmean=7, psd=2, pdist='gaussian')  
setMosqBitingPrior(pmean=0.25, psd=0.05, pdist='gaussian')  
setHumanLifeExpPrior(pmean=73, psd=2, pdist='gaussian')
setHumanIncPerPrior(pmean=5, psd=1, pdist='gaussian')
setHumanInfPerPrior(pmean=5, psd=1, pdist='gaussian')
setHumanMosqTransProbPrior(pmean=0.5, psd=0.01, pdist='gaussian')

estimateEcoCoefficients(nMCMC=100000, bMCMC=0.5, cRho=0.1, cEta=1, gauJump=0.75)
simulateEmpiricalIndexP(nSample=1000, smoothing=c(7,15,30,60))

exportEmpiricalIndexP()
plotEmpiricalIndexP(outfilename='debug_indexP')


#processing RJ indexP output into weekly indexP####
require(tidyverse)
require(dplyr)
indexP_RJ <- read.csv("RJ/.estimated_indexP.csv", header = TRUE)
indexP_RJ_weekly <- indexP_RJ %>%
  select(date, indexP) %>%
  mutate(date = as.Date(date),
         daily_indexP = indexP) %>%
  group_by(week_start_date = lubridate::floor_date(date, "week", week_start = getOption("lubridate.week.start", 3))) %>% ##week starts from 1st Jan 2014
  mutate(weekly_indexP = mean(indexP))

ggplot(indexP_RJ_weekly, aes(x = week_start_date, y = weekly_indexP))+
  geom_point()

#output weekly data and save to csv
indexP_RJ_weekly_out <- indexP_RJ_weekly %>%
  select(week_start_date, weekly_indexP) %>%
  distinct(week_start_date, .keep_all = TRUE)

#write out the daily indexP and weekly
write.csv(indexP_RJ_weekly, file = "RJ/RJ_daily_weekly_indexP.csv", row.names = FALSE)

#write out the mean weekly indexP
write.csv(indexP_RJ_weekly_out, file = "RJ/RJ_weekly_indexP.csv", row.names = FALSE)
```
