## Supplementary File S1 for "Epidemiological and genomic investigation of chikungunya virus in Rio de Janeiro state, Brazil, between 2015 and 2018": Estimate_R0.R

```
#Load packages
library(lubridate)
library(dplyr)
library(R0)

# Data
dates <- c("2016-01-03",
           "2016-01-10",
           "2016-01-17",
           "2016-01-24",
           "2016-01-31",
           "2016-02-07",
           "2016-02-14",
           "2016-02-21",
           "2016-02-28",
           "2016-03-06",
           "2016-03-13")
values <- c(25,41,44,54,78,68,101,99, 202,210,287)

# adjust data
epid <- data.frame(cbind(dates,values))
names(epid) <- c("t","incid")
epid$incid <- as.numeric(epid$incid)
epid$t <- as.Date(epid$t)

# Run regression
tmp <- lm((log(incid)) ~ t, data=epid)

# get statistics
Rsquared = summary(tmp)$r.squared
r <- coefficients(tmp)[2]
conf.int = confint(tmp)[2,]

# Generation time distribution for CHIKV: https://www.pnas.org/doi/epdf/10.1073/pnas.1611391113
mean_si <- 14 # days
std_si <- sqrt(41) # days
mGT<-generation.time("gamma", c(mean_si, std_si))
R = as.numeric(R.from.r(r,mGT))
R.inf = as.numeric(R.from.r(conf.int[1],mGT))
R.sup = as.numeric(R.from.r(conf.int[2],mGT))
paste(R,R.inf,R.sup)

# Sensitivity 10 days
mean_si <- 10 # days
std_si <- sqrt(41) # days
mGT<-generation.time("gamma", c(mean_si, std_si))
R = as.numeric(R.from.r(r,mGT))
R.inf = as.numeric(R.from.r(conf.int[1],mGT))
R.sup = as.numeric(R.from.r(conf.int[2],mGT))
paste(R,R.inf,R.sup)

# Sensitivity 20 days
mean_si <- 20 # days
std_si <- sqrt(41) # days
mGT<-generation.time("gamma", c(mean_si, std_si))
R = as.numeric(R.from.r(r,mGT))
R.inf = as.numeric(R.from.r(conf.int[1],mGT))
R.sup = as.numeric(R.from.r(conf.int[2],mGT))
paste(R,R.inf,R.sup)

###############################################################################
# R0 estimates With genetic data
# r was obtained from a beast estimate
r = 5.159/365 #days
r.inf = 0.7935/365 # days
r.sup = 9.7395/365 # days


# Real
mean_si <- 14 # days
std_si <- sqrt(41) # days
mGT<-generation.time("gamma", c(mean_si, std_si))
R = as.numeric(R.from.r(r,mGT))
R.inf = as.numeric(R.from.r(r.inf,mGT))
R.sup = as.numeric(R.from.r(r.sup,mGT))
paste(R,R.inf,R.sup)

# Sensitivity 10 days
mean_si <- 10 # days
std_si <- sqrt(41) # days
mGT<-generation.time("gamma", c(mean_si, std_si))
R = as.numeric(R.from.r(r,mGT))
R.inf = as.numeric(R.from.r(r.inf,mGT))
R.sup = as.numeric(R.from.r(r.sup,mGT))
paste(R,R.inf,R.sup)

# Sensitivity 20 days
mean_si <- 20 # days
std_si <- sqrt(41) # days
mGT<-generation.time("gamma", c(mean_si, std_si))
R = as.numeric(R.from.r(r,mGT))
R.inf = as.numeric(R.from.r(r.inf,mGT))
R.sup = as.numeric(R.from.r(r.sup,mGT))
paste(R,R.inf,R.sup)
```
