## Supplementary File S1 for "Epidemiological and genomic investigation of chikungunya virus in Rio de Janeiro state, Brazil, between 2015 and 2018": Estimate_Rt.R

```
# Estimate Rt for CHIKV

# Load libraries
library(lubridate)
library(EpiEstim)
library(ggplot2)
library(incidence)
library(cowplot)

# Load incidence data
df <- read.csv("number_of_chikv_confirmed_cases_RJ.csv")
head(df)

# Convert dates
df$date <- dmy(df$date)
head(df)

# Subset the dataframe and keep only data from 2016
df <- df[c("date","value")]
df <- df[which(df$date > dmy(01012016)),]


# Serial interval for chikv: https://www.pnas.org/doi/epdf/10.1073/pnas.1611391113
mean_si <- 14 # days
std_si <- sqrt(41) # days
mean_si <- mean_si/7 # weeks
std_si <- std_si/7 # weeks

T <- nrow(df)
t_start <- seq(2, T-7) # starting at 2 as conditional on the past observations
t_end <- t_start + 7   # 8 weeks windows for weekly incidence data

# Estimate Rt with EpiEstim
res <- estimate_R(df$value, 
                  method="parametric_si",
                  config = make_config(list(
                    t_start = t_start,
                    t_end = t_end,
                    mean_si = mean_si, 
                    std_si = std_si))
)
#plot(res,legend = F)
plot(res,"R",legend = F)


# Sensitivity analysis - 10
mean_si <- 10 # days
std_si <- sqrt(41) # days
mean_si <- mean_si/7 # weeks
std_si <- std_si/7 # weeks

# Estimate Rt with EpiEstim
res <- estimate_R(df$value, 
                  method="parametric_si",
                  config = make_config(list(
                    t_start = t_start,
                    t_end = t_end,
                    mean_si = mean_si, 
                    std_si = std_si)))
#plot(res,legend = F)
plot(res,"R",legend = F)


# Sensitivity analysis - 20
mean_si <- 20 # days
std_si <- sqrt(41) # days
mean_si <- mean_si/7 # weeks
std_si <- std_si/7 # weeks

# Estimate Rt with EpiEstim
res <- estimate_R(df$value, 
                  method="parametric_si",
                  config = make_config(list(
                    t_start = t_start,
                    t_end = t_end,
                    mean_si = mean_si, 
                    std_si = std_si)))
#plot(res,legend = F)
plot(res,"R",legend = F)
```
